# Multimodal Machine Learning for Diagnosis of Multiple Sclerosis Using Optical Coherence Tomography in Pediatric Cases

**DOI:** 10.1101/2025.09.12.25335649

**Authors:** Chaojun Chen, Sahar Soltanieh, Sajith Rajapaksa, Farzad Khalvati, Ann Yeh

## Abstract

**Background and Objectives:** Identifying MS in children early and distinguishing it from other neuroinflammatory conditions of childhood is critical, as early therapeutic intervention can improve outcomes. The anterior visual pathway has been demonstrated to be of central importance in diagnostic considerations for MS and has recently been identified as a fifth topography in the McDonald Diagnostic Criteria for MS. Optical coherence tomography (OCT) provides high-resolution retinal imaging and reflects the structural integrity of the retinal nerve fiber and ganglion cell inner plexiform layers. Whether multimodal deep learning models can use OCT alone to diagnose pediatric MS (POMS) is unknown.

**Methods:** We analyzed 3D OCT scans collected prospectively through the Neuroinflammatory Registry of the Hospital for Sick Children (REB#**1000005356**). Raw macular and optic nerve head images, and 52 automatically segmented features were included. We evaluated three classification approaches: (1) deep learning models (e.g. ResNet, DenseNet) for representation learning followed by classical ML classifiers, (2) ML models trained on OCT-derived features, and (3) multimodal models combining both via early and late fusion.

**Results:** Scans from individuals with POMS (onset **16.0 *±* 3.1** years, **51.0%**F; **211** scans) and **29** children with non-inflammatory neurological conditions (**13.1 *±* 4.0** years, **69.0%**F, **52** scans) were included. The early fusion model achieved the highest performance (AUC: **0.87**, F1: **0.87**, Accuracy: **90%**), outperforming both unimodal and late fusion models. The best unimodal feature-based model (SVC) yielded an AUC of **0.84**, F1 of **0.85** and an accuracy of **85%**, while the best image-based model (ResNet101 with Random Forest) achieved an AUC of **0.87**, F1 of **0.79**, and accuracy of **84%**. Late fusion underperformed, reaching **82%** accuracy but failing in the minority class.

**Discussion:** Multimodal learning with early fusion significantly enhances diagnostic performance by combining spatial retinal information with clinically relevant structural features. This approach captures complementary patterns associated with MS pathology and shows promise as an AI-driven tool to support pediatric neuroinflammatory diagnosis.

## 1 Introduction

Multiple sclerosis (MS) is a chronic, immune-mediated disease of the central nervous system that most commonly presents in adults between the ages of 20 and 40 [1]. Pediatric-onset multiple sclerosis (POMS) accounts for approximately 3–5% of all MS cases [2]. Early and accurate diagnosis is critical, as neurodegeneration begins at disease onset, and timely initiation of disease-modifying therapies can significantly improve long-term outcomes [3]. In children with MS, the optic nerve is often among the earliest sites of inflammation [4–6], which has led to increased clinical attention to the visual pathway as a diagnostic target.

Reflecting this emphasis, recent revisions to the McDonald diagnostic criteria have highlighted the optic nerve as a key topographic site, recommending the use of imaging modalities such as magnetic resonance imaging (MRI), optical coherence tomography (OCT), and visual evoked potentials (VEP) to establish dissemination in space [7]. Yet, despite improved imaging protocols, diagnosis in pediatric MS remains more ambiguous than in adults, due to overlapping features with other inflammatory and genetic conditions.

Among diagnostic modalities relevant to MS, OCT has emerged as a powerful, non-invasive imaging tool capable of capturing high-resolution structural details of the retina, including the retinal nerve fiber layer (RNFL) and ganglion cell layer (GCL)—regions frequently affected in MS. OCT-derived features have been shown to correlate with disease severity and visual dysfunction, with demonstrated utility in MS diagnosis and monitoring [5, 8–14]. Parallel to these developments, machine learning (ML) has gained traction in medical imaging, showing strong potential in disease detection [15–17], medical image analysis [18, 19], patient outcome prediction [20, 21], and personalized treatment planning [22, 23].

Despite this progress, most ML applications in MS have remained unimodal, relying on a single data source such as imaging [24–26], electronic health records [27–29], or genomic sequences [30, 31]. While these approaches have provided useful insights, they fail to capture the multifaceted nature of MS, particularly in pediatric cases where diagnostic uncertainty is greater. In addition, clinical diagnosis often involves integrating information from multiple modalities, and unimodal ML models may overlook important cross-modal interactions. Moreover, most existing studies compare MS patients to healthy controls, a simplification that does not reflect the real-world clinical scenario. Few studies have investigated OCT-based ML for pediatric MS, and fewer still use diagnostically challenging comparator groups.

Multimodal ML, which integrates heterogeneous data sources to improve model robustness and generalizability may address some of these limitations [32–34]. In the context of OCT, combining raw 3D image volumes with extracted structural features provides a complementary perspective that may enhance disease modeling and classification accuracy.

In this study, we perform a comprehensive evaluation of unimodal and multimodal ML models for classifying pediatric MS using OCT data. Importantly, our comparator group consists not of healthy controls, but children with non-inflammatory neurological conditions, many of whom present with overlapping symptoms and abnormal imaging, making the classification task more clinically realistic and challenging. Our study assesses deep learning classifiers trained on raw 3D OCT volumes, classical ML models trained on quantitative OCT-derived features, and two multimodal integration strategies, early fusion and late fusion. Our benchmarking framework compares classification performance across these configurations, analyzing the effects of model architecture, feature representation, and fusion strategy on accuracy, robustness, and class sensitivity. This work represents one of the first detailed investigations into multimodal ML for pediatric MS using OCT and establishes a foundation for future clinical applications in neuroinflammatory disorders.

## 2 Methods

### 2.1 Participants and Eligibility Criteria

This study utilizes consecutive OCT scans performed on children and youth as part of an ongoing prospective registry (Neuroinflammatory Registry) at the Hospital for Sick Children, Toronto, Canada (Research Ethics Board protocol #1000005356). Informed consent and assent were obtained from all participants prior to data collection.

We included OCTs from individuals diagnosed with MS and from children with other non-inflammatory neurological diagnoses. To ensure data quality, diagnostic accuracy, and consistency across participant groups, we applied the following inclusion and exclusion criteria during dataset curation, as summarized in Figure 1.

**Fig. 1.**
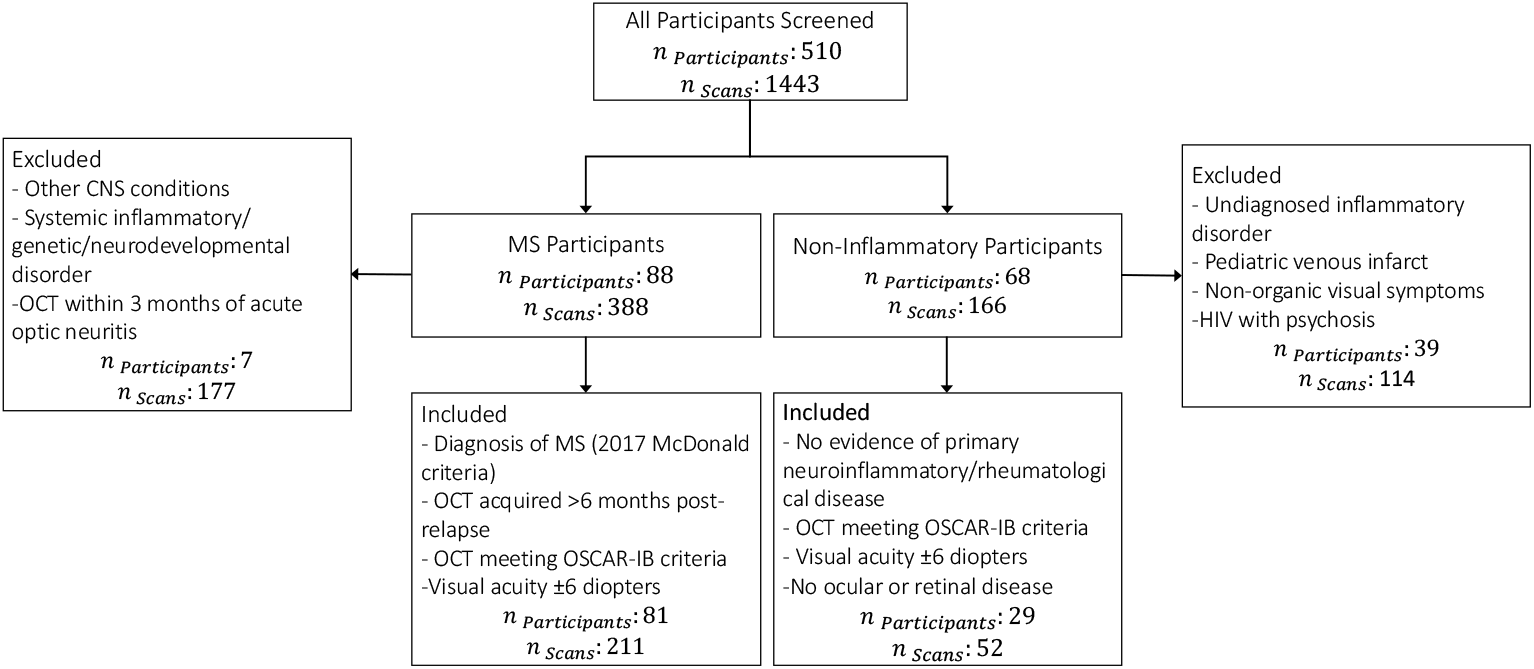
Flowchart illustrating the inclusion and exclusion criteria applied for participant selection in the study. Individuals with confirmed MS diagnoses and non-inflammatory neurological conditions were screened based on clinical, imaging, and quality control parameters to ensure consistency and data integrity.

- Inclusion criteria for MS participants:

– Confirmed diagnosis of MS according to the 2017 McDonald criteria [7]
– OCT acquired more than 6 months after an acute relapse
– OCT meeting OSCAR-IB quality control criteria [35]
– Visual acuity within ± 6 diopters

- Exclusion Criteria for MS participants:

– Inability to tolerate OCT testing
– Presence of other central nervous system (CNS) conditions
– Diagnosis of concomitant systemic inflammatory, genetic, or neurodevelopmental disorders
– OCT acquired within 3 months of an acute optic neuritis episode, to avoid confounding due to retinal swelling

- Inclusion criteria for Non-Inflammatory participants:

– No clinical or laboratory evidence of a primary acquired neuroinflammatory or rheumatological disease, based on serum and cerebrospinal fluid (CSF) assessments.
– High-quality OCT scans meeting OSCAR-IB criteria [35]
– Visual acuity within the ± 6 diopters.
– No evidence for ocular or retinal disease.

- Exclusion criteria for Non-Inflammatory participants:

– Inability to tolerate OCT testing.
– OCT scans or clinical history suggestive of conditions that could confound interpretation, including:

∗ Possible undiagnosed inflammatory disorder
∗ Pediatric venous infarct with stable white matter abnormalities
∗ Functional visual symptoms associated with anxiety
∗ Virally suppressed HIV with comorbid psychosis

The non-inflammatory cohort comprised individuals with diverse neurological and systemic conditions (Supplementary Table 1).

To support robust model development and generalizability, the dataset is partitioned into training and testing subsets:

- **Training Set:** Includes multiple OCT scans per patient across different visits to capture longitudinal variability. It consists 27 male, 40 female, and 2 unknown participants (mean age 13.77 ± 3.23 years), comprising 179 scans from MS patients and 43 from controls.
- **Testing Set:** Limited to the first-visit scan per participant to ensure an unbiased evaluation, with scans obtained at least 6 months after an episode of optic neuritis. It includes 11 male, 29 female, and 1 unknown participant (mean age 16.29 ± 3.49 years), with 32 MS and 9 control scans.

Each OCT scan included two data modalities: (1) 3D OCT Volumes, which are raw volumetric images covering a 6 × 6 × 2*mm*^3^ region centered on the macula, and (2) autosegmented OCT features, comprising 52 quantitative measures per scan, including layer-wise thickness values for the RNFL and GCIPL. A detailed breakdown of the demographics of the participants and the distributions of the diagnostic groups is provided in Table 1.

**Table 1.** Summary of dataset demographics of the studied population. P values compare MS and Non-inflammatory groups using the Chi-squared test for categorical variables and Mann–Whitney U test for continuous variables.

| Variable | Total<br>(*n* = 110) | MS Patients<br>(*n* = 81) | Non-inflammatory<br>(*n* = 29) | P value |
| --- | --- | --- | --- | --- |
| **Number of Scans** | 263 | 211 | 52 | – |
| **Sex, n (%):** |  |  |  |  |
| Female | 71 (64.5%) | 51 (63.0%) | 20 (69.0%) | 0.6126 |
| **Age (mean ± SD)** | 15.3 ± 3.6 | 16.0 ± 3.1 | 13.1 ± 4.0 | 0.0023 |
| **Age (Range)** | 5 – 18 | 9 – 18 | 5 – 18 | – |

### 2.2 OCT Acquisition Protocol

OCT imaging was performed using a spectral-domain Cirrus HD-OCT scanner (Model 5000, Carl Zeiss Meditec). All scans were acquired under standardized clinical conditions by experienced technicians trained in pediatric OCT imaging. For each participant, two types of scans were obtained: (1) Macular Cube (512 × 128), which captures a 6 × 6 × 2 *mm*^3^ volume centered on the macula, and (2) Optic Nerve Head Cube (200 × 200), which captures a comparable volume centered on the optic nerve head. In this study, we select the Macular Cube scans, as they more consistently reflect neurodegenerative changes associated with multiple sclerosis (MS), particularly thinning in the ganglion cell layer and inner plexiform layer, recognized biomarkers of MS even in individuals without a history of optic neuritis.

All OCT scans underwent rigorous quality control using the OSCAR-IB criteria, [35] adapted for the Cirrus HD-OCT platform. These criteria include: O − Absence of acquisition artifacts, S – Signal strength ≥ 7, C – Correct centration, A – No segmentation failures, R – Absence of retinal pathology, I – Adequate fundus illumination, B – Proper beam placement.

Feature extraction was performed using the device’s automated segmentation software. RNFL thickness is analyzed both globally and across four quadrants: superior, inferior, nasal, and temporal. GCIPL thickness is represented by eight features: average, minimum, and six sectoral measurements, superotemporal, superior, superonasal, inferonasal, inferior, and inferotemporal. These features were selected based on their established clinical relevance in identifying MS-associated neurodegeneration, as demonstrated in prior studies. [36] A complete list of the extracted OCT features used in this study is provided in Appendix 2.

Table 2 presents the average and standard deviation of selected OCT-derived parameters for both MS and non-inflammatory groups. These include global and regional retinal layer thicknesses, foveal and ganglion cell layer measurements, as well as clinically relevant inter-eye differences and RNFL thinning metrics commonly associated with neurodegeneration in MS. To compute these statistics, we include only the first available (baseline) scan per patient.

**Table 2.** Summary of OCT features for MS and Non-Inflammatory Groups. Values are reported as ± mean standard deviation or count (percentage) as appropriate.

| *OCT Parameter / Statistic* | *MS*<br>(*n* = 211) | *Non-Inflammatory*<br>(*n* = 52) |
| --- | --- | --- |
| **Retinal Layer Thickness (in $\mu m$)** |  |  |
| RNFL Thickness | 93.55±13.85 | 98.53±6.30 |
| Superior Quadrant Thickness | 121.59±18.88 | 127.34±10.58 |
| Nasal Quadrant Thickness | 70.72±12.13 | 72.72±10.70 |
| Inferior Quadrant Thickness | 121.81±22.70 | 126.14±12.65 |
| Temporal Quadrant Thickness | 60.05±15.01 | 68.34±9.98 |
| Foveal Thickness (Inner) | 253.44±7.85 | 253.55±10.94 |
| Foveal Thickness (Outer) | 66.33±3.43 | 67.22±2.85 |
| Ganglion Cell Layer Thickness (GCL) | 76.76±9.48 | 85.34±4.16 |
| **Clinical Threshold-Based Metrics** |  |  |
| Inter-Eye RNFL Difference ($\mu m$) | 8.84±15.47 | 3.34±2.79 |
| # Inter-Eye RNFL Difference ≥ 6 | 29 (35%) | 4 (13%) |
| # RNFL Thickness ≤ 87 | 23 (28%) | 2 (6%) |

### 2.3 Data Preprocessing

To prepare the OCT data for model training, a series of preprocessing steps was applied to the raw 3D OCT volumes acquired independently from the left and right eyes of each subject. An overview of the full preprocessing pipeline is illustrated in Figure 2. Each 3D volume was first spatially downsampled by 50% along the height and width dimensions using a zoom factor of (1, 0.5, 0.5), where the dimensions correspond to (depth, height, width), to reduce computational overhead while preserving anatomical content.

**Fig. 2.**
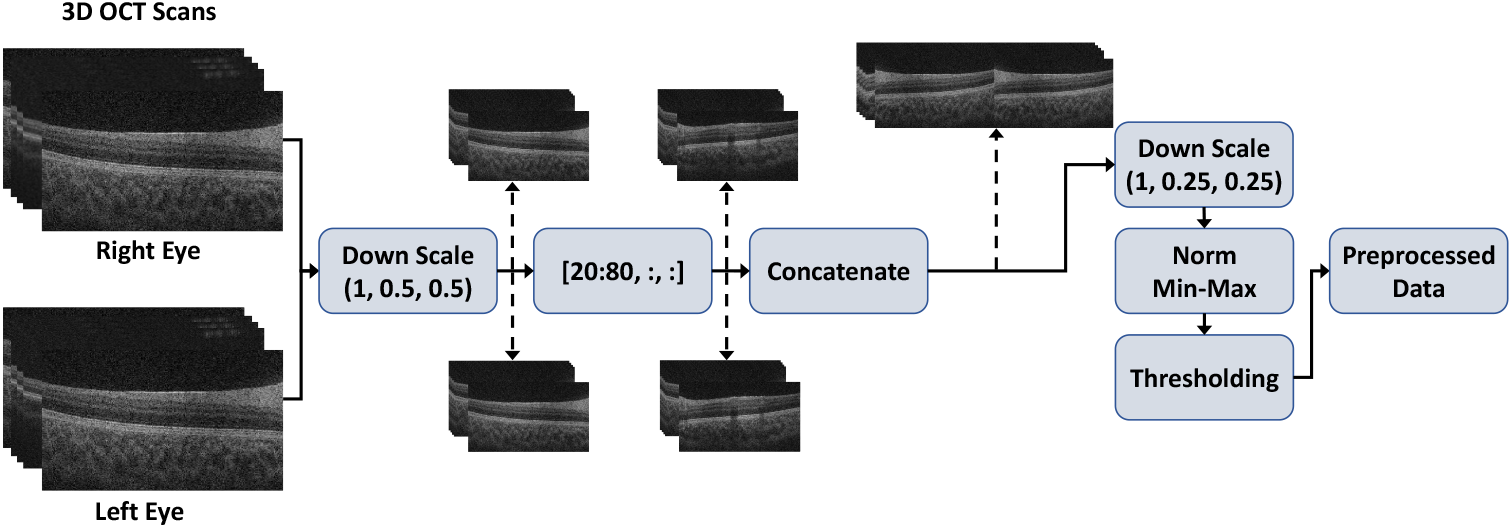
Overview of the preprocessing pipeline.

Next, only the central slices, specifically slices 20 to 80, were retained from each volume. This selection focuses the analysis on the most informative retinal regions. The volumes from the left and right eyes were then concatenated along the width axis, effectively placing them side by side. This resulted in a single unified volume where the height and depth remained unchanged, and the width became twice that of each individual eye volume.

To further reduce the spatial resolution and memory footprint, an additional down-sampling step was performed using a zoom factor of (1, 0.25, 0.25). The concatenated volumes were then normalized using min-max normalization, where each voxel intensity was scaled based on the following formula:

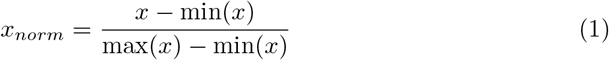

where *x*_*norm*_ denotes the normalized voxel intensity values, and max(*x*) and min(*x*) represent the maximum and minimum intensity values within the volume, respectively. Finally, a thresholding operation is applied to suppress low-intensity background noise. All voxel intensities below a threshold of 0.3 are set to zero, while higher intensities are retained. This operation is defined as:

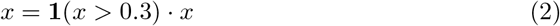

where **1** is an indicator function that returns 1 if the condition is satisfied and 0 otherwise.

This preprocessing pipeline ensured that the input data retained relevant anatomical features while remaining computationally efficient for deep learning model training.

### 2.4 Model Architectures and Fusion Strategies

We explored both unimodal and multimodal strategies for OCT-based classification, leveraging deep learning and classical machine learning methods.

#### 2.4.1 Unimodal Approaches

The unimodal setting involved training separate models on two individual modalities: volumetric 3D OCT scans, and (2) a set of quantitative OCT-derived features.

##### 3D OCT Scans

For raw volumetric 3D OCT data, we investigated two modeling strategies:

- **End-to-End Deep Learning Models:** We trained several 3D convolutional neural networks to directly learn from OCT volumes. The architectures evaluated include: DenseNet, ResNet34, ResNet50, and ResNet110.

– **3D ResNet:** ResNet employs identity-based skip connections to mitigate vanishing gradients in deep networks. We evaluate three variants, including ResNet34, ResNet50, and ResNet101, each composed of a stack of residual blocks. Each block consists of 3D convolutions, batch normalization, and ReLU activations, followed by an identity connection that adds the input to the output. The input 3D OCT volume passes through an initial 3D convolution and max-pooling layer, followed by residual blocks and global average pooling. A fully connected layer is used for classification.
– **3D DenseNet:** DenseNet introduces dense connectivity, where each layer receives inputs from all preceding layers and passes its output to all subsequent layers. This design promotes feature reuse and mitigates the vanishing gradient problem. Our implementation includes multiple dense blocks containing 3D convolutional layers, interleaved with transition layers that use 3D pooling and 1 × 1 × 1 convolutions for dimensionality reduction. Final features are globally pooled and fed into a fully connected classification layer.

- **Hybrid Representation + Classifier Models:** We extracted feature representations from the last convolutional layer of the trained CNNs and use them as input to classical machine learning classifiers. The classifiers include:

– **Random Forest:** [37] An ensemble of decision trees with *n estimators* = 10, using class weights of 0 : 1, 1 : 3 to address class imbalance.
– **Gradient Boosting** [38]: A sequential ensemble method trained with *n estimators* = 10.
– **XGBoost:** [39] An optimized gradient boosting algorithm using *n estimators* = 10 and *scale pos weight* = 3 to compensate for class imbalance.
– **K-Nearest Neighbors (KNN):** [40] A non-parametric method with *n neighbors* = 20, classifying based on the majority label among nearest neighbors.
– **Support Vector Classifier (SVC):** [41] A linear SVM with *kernel* =^*′*^ *linear*^*′*^, *probability* = *True*, and class weights of 0 : 1, 1 : 3.
– **Fully Connected Neural Network (NN):** [42] A multilayer perceptron with two hidden layers of 100 and 50 units, ReLU activation, and Adam optimizer, trained for up to 500 iterations.

##### Quantitative OCT Features

We also trained models on a set of pre-extracted quantitative OCT features. Prior to classification, we apply Least Absolute Shrinkage and Selection Operator (LASSO) regression to select the top six most informative features. These selected features are used to train the same set of classical classifiers described above.

#### 2.4.2 Multimodal Approaches

In the multimodal setting, we integrated information from both raw 3D OCT scans and OCT-derived features using two fusion strategies: early fusion and late fusion.

- **Early Fusion**

For early fusion, we first extracted feature-level representations from each modality:

– Deep CNN-derived feature vectors from the 3D OCT scans (ResNet and DenseNet)
– The full set of quantitative OCT features

These vectors were concatenated and standardized using StandardScaler. To reduce dimensionality and enhance interpretability, we applied LASSO to the combined representation, selecting the top six features across both modalities. The resulting vector is then used to train the same set of classical classifiers: Random Forest, Gradient Boosting, XGBoost, KNN, SVC, and NN. The pipeline is shown in Figure 3.

**Fig. 3.**
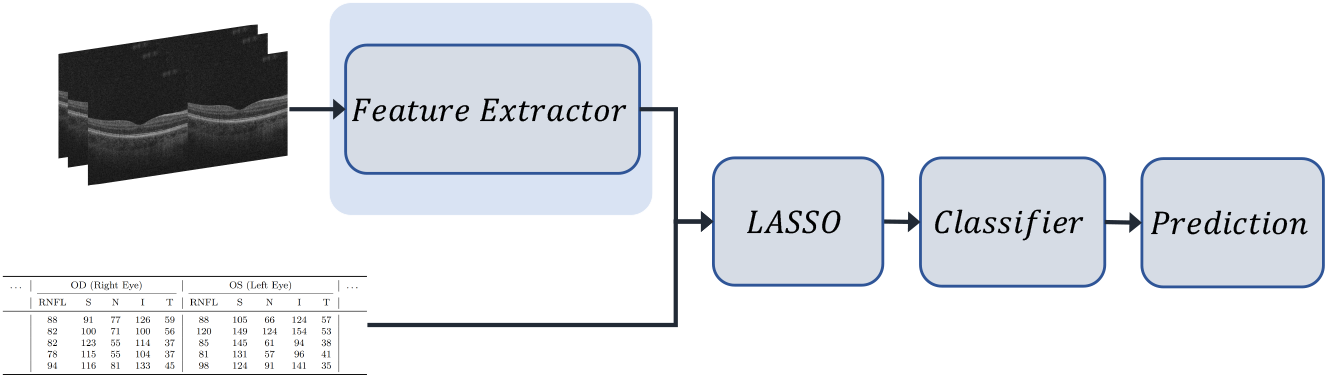
Overview of the early fusion pipeline for MS classification. Deep features are first extracted from 3D OCT volumes using a CNN-based model. These image-derived embeddings are concatenated with tabular OCT-derived clinical features. The combined feature vector is then passed through LASSO for dimensionality reduction and used as input to a classifier for final prediction.

- **Late Fusion**

We took the best-performing unimodal classifiers from each modality and averaged their predicted probabilities to produce the final prediction, as shown in Figure 4 This ensemble strategy leveraged the complementary strengths of each modality, enhancing robustness and generalization.

### 2.5 Implementation Details

All experiments were implemented in Python 3.9.13 using the PyTorch deep learning framework. Key libraries include NumPy for numerical operations, scikit-learn for evaluation metrics and data handling, and SciPy for scientific computing. A fixed *random state* = 50 is used across all scikit-learn models to ensure reproducibility. Model training is performed on a single NVIDIA Tesla V100 GPU (32 GB VRAM) with CUDA version 12.8. The 3D OCT models are trained using the AdamW optimizer with a learning rate of 0.001 and a batch size of 4. Training runs for 30 epochs using binary cross-entropy loss (nn.BCELoss()), and a dropout rate of 0.1 is applied for regularization.

**Fig. 4.**
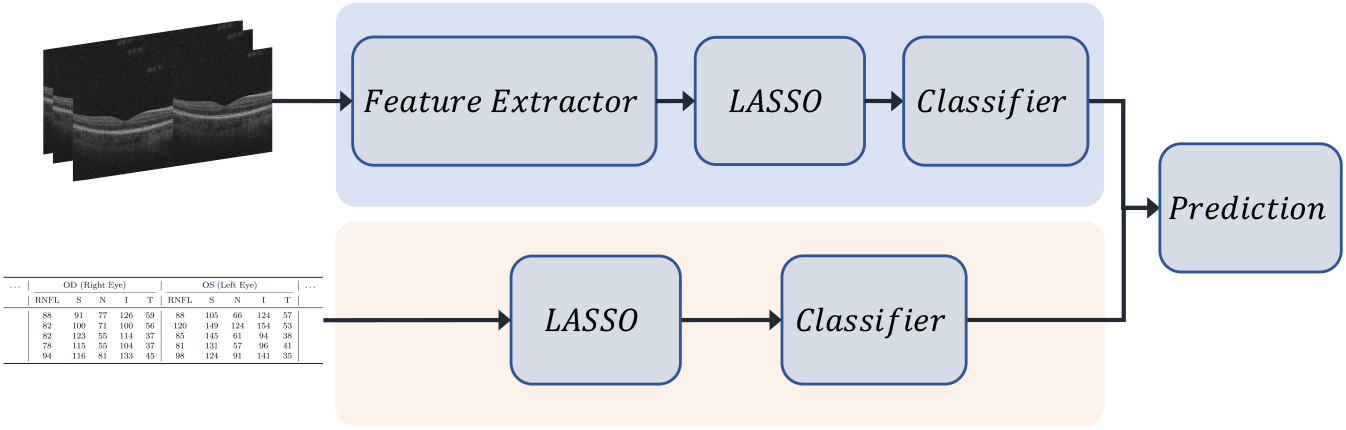
Overview of the late fusion pipeline. Separate classifiers are trained independently on 3D OCT image-derived features and tabular OCT-derived clinical features. Each classifier outputs a probability distribution over the target classes. Final predictions are obtained by averaging the output probabilities from both classifiers.

### 2.6 Evaluation Protocol

To ensure robust model selection while avoiding data leakage, we adopted a 5-fold cross-validation strategy on the training set. The training data was divided into five stratified folds at the patient level, ensuring that each fold maintained a representative class distribution and that no patient appeared in more than one fold. In each iteration, four folds are used for training and one for validation. This process was repeated five times, allowing each fold to serve once as the validation set. The cross-validation results guided the selection of the best-performing model.

After model selection, we evaluated the final model on a held-out test set that remains untouched during both training and validation. We report three evaluation metrics: Accuracy, F1 score, and Area Under the Receiver Operating Characteristic Curve (AUROC).

To account for class imbalance, we also report the F1 score, which calculates the F1 score for each class and averages them using the number of true instances for each class as weights. This ensures that classes with more samples contribute proportionally more to the final score, making it particularly appropriate for imbalanced datasets. AUROC quantifies the model’s ability to discriminate between the positive and negative classes across all classification thresholds. It is computed as the area under the Receiver Operating Characteristic (ROC) curve, which plots the True Positive Rate (TPR) against the False Positive Rate (FPR) at various thresholds.

AUROC is a threshold-independent metric and is especially useful in imbalanced settings. By reporting AUROC alongside Accuracy and F1 score, we provide a comprehensive evaluation of the model’s performance, capturing both its discrimination capability and classification effectiveness.

## 3 Results

This section presents a comprehensive evaluation of the proposed models and fusion strategies for MS classification. We begin by analyzing the performance of uni-modal approaches, separately assessing models trained solely on volumetric OCT images and those using tabular OCT features. Subsequently, we investigate multi-modal fusion techniques designed to leverage complementary information from both data types, employing early fusion with feature concatenation and dimensionality reduction, as well as late fusion through ensemble averaging of modality-specific classifiers.

### 3.1 Unimodal Classification with 3D OCT Volumes

We first evaluate unimodal classification using volumetric 3D OCT data. The results of top 5 performing models are presented in Table 3.

**Table 3.** Top 5 performing models for 3D OCT images classification. The number in parentheses next to each feature extractor model indicates the dimensionality of the extracted features used for classification

| Feature Extractor → Classifier | F1 Score | AUC | Accuracy |
| --- | --- | --- | --- |
| ResNet101 (256) → Random Forest | 0.79 | 0.87 | 0.84 |
| DenseNet (256) → Random Forest | 0.65 | 0.87 | 0.86 |
| ResNet101 (256) → XGBoost | 0.75 | 0.82 | 0.80 |
| DenseNet (2048) → NN | 0.71 | 0.85 | 0.80 |
| DenseNet (128) → NN | 0.67 | 0.85 | 0.80 |

The best-performing configuration is ResNet101 with 256-dimensional embeddings, followed by Random Forest, achieving an F1 score of 0.79, an AUC of 0.87, and an accuracy of 0.84. Larger embeddings (e.g., 2048) tended to degrade performance due to overfitting, especially in margin-based classifiers like SVC.

KNN classifiers showed mixed results. In some configurations, such as DenseNet128, performance is strong (*F* 1 = 0.7951), but overall, KNN tended to struggle with generalization. This behavior is likely related to its sensitivity to data sparsity and the curse of dimensionality, which affect the reliability of distance-based metrics in higher-dimensional spaces.

XGBoost classifiers produced moderately competitive results in a few configurations. For instance, ResNet101 with 256-dimensional features, achieved an F1 score of 0.75, AUC of 0.82, and an accuracy of 80%. Although looking at the trend, they are less consistent compared to Random Forest models, which more robustly handle variance in feature distributions.

DenseNet-based models paired with ensemble classifiers such as Random Forest showed competitive performance. Notably, several models achieved perfect specificity (no false positives), although this often occurs at the expense of reduced sensitivity, particularly in KNN and SVC classifiers.

An important observation is that intermediate-dimensional feature embeddings (e.g., dimensions of 128 or 256) often outperform higher-dimensional ones (e.g., 2048), particularly when combined with tree-based classifiers. This trend suggests that high-dimensional representations may introduce noise or increase the risk of overfitting if not properly regularized.

### 3.2 Unimodal Classification with Tabular OCT Features

In this section, we conduct an extensive evaluation of the performance of different models in the classification task using the OCT features. Among the classifiers, the Support Vector Classifier (SVC) achieved the highest AUC (0.84), accuracy (84.62%), and weighted F1 score (0.85), demonstrating superior performance in distinguishing between classes. It outperformed traditional ensemble methods such as Random Forest (AUC = 0.70) and XGBoost (AUC = 0.71), and significantly outperformed NN and KNN as shown in Table 4. This suggests that a linear decision boundary with appropriate class balancing is most effective given the selected OCT features.

**Table 4.** Performance of OCT feature-based models.

| Model | F1 Score | AUC | Accuracy |
| --- | --- | --- | --- |
| Random Forest | 0.81 | 0.70 | 0.82 |
| Gradient Boosting | 0.78 | 0.62 | 0.82 |
| KNN | 0.78 | 0.65 | 0.79 |
| SVC | **0.85** | **0.84** | **0.85** |
| XGBoost | 0.75 | 0.71 | 0.74 |
| NN | 0.74 | 0.64 | 0.82 |

The models’ performances indicate that while some models, such as NN, predicted only the majority class (sensitivity = 0 for the minority class), SVC maintained a more balanced classification with 57% correct positive predictions.

### 3.3 Early Fusion: Feature-Level Integration

To evaluate the benefit of incorporating clinical data alongside 3D OCT volumes, we implemented an early fusion strategy. DenseNet feature extractor with a 128-dimensional embedding, followed by an SVC classifier, achieved the best performance (AUC = 0.90, Accuracy = 87.18%, F1 = 0.87). The results for this configuration show a true negative rate of 90%, a true positive rate of 71%, a false positive rate of 9%, and a false negative rate of 28%.

Multiple CNN architectures demonstrated robust performance under the multimodal framework. ResNet101 (128-dimensional), ResNet34 (512-dimensional), and ResNet50 (2048-dimensional), all combined with SVC classifiers, achieved identical accuracy and F1 scores of 87.18% and 0.87, respectively. These results suggest that different CNN-based extractors can yield similarly high performance when fused with clinical data and appropriately regularized through LASSO. Top-performing model results are presented in Table 5.

**Table 5.** Top-performing multimodal classification models (Early Fusion + LASSO). The CNN output dimensionality refers to the size of the last linear layer in the feature extractor.

| Feature Extractor → Classifier | F1 Score | AUC | Accuracy |
| --- | --- | --- | --- |
| DenseNet (128) → SVC | 0.87 | 0.90 | 0.87 |
| ResNet101 (128) → SVC | 0.87 | 0.87 | 0.87 |
| ResNet34 (512) → SVC | 0.87 | 0.87 | 0.87 |
| ResNet50 (2048) → SVC | 0.87 | 0.86 | 0.87 |
| ResNet101 (2048) → SVC | 0.85 | 0.85 | 0.85 |
| ResNet101 (256) → SVC | 0.85 | 0.85 | 0.85 |
| ResNet34 (256) → SVC | 0.85 | 0.85 | 0.85 |
| ResNet50 (128) → SVC | 0.84 | 0.85 | 0.84 |
| ResNet50 (256) → SVC | 0.84 | 0.85 | 0.84 |
| DenseNet (256) → SVC | 0.79 | 0.85 | 0.81 |

### 3.4 Late Fusion: Decision-Level Integration

Late fusion combined the output probabilities from the best unimodal classifiers: ResNet101 (256) + Random Forest (3D) and SVC (tabular). The averaged output yielded AUC = 0.7902, accuracy = 82.05%, F1 = 0.73. This result reflects perfect classification of negatives but poor performance in identifying positives.

This underperformance illustrates the limitations of late fusion, especially its inability to model cross-modal interactions and compensate for weak classifiers in one modality. To summarize performance across different input modalities and fusion strategies, we compare the best-performing models from each experimental setting in Table 6. The results clearly indicate that early fusion achieves superior performance across all three evaluation metrics, F1 score, AUC, and accuracy, demonstrating the value of jointly modeling clinical and imaging data. In contrast, late fusion, which combines independent predictions at the decision level, underperforms significantly, failing to correctly identify any positive samples despite high accuracy. This discrepancy underscores the limitations of post hoc ensemble averaging in handling class imbalance and capturing cross-modal interactions. Notably, the unimodal tabular approach using SVC performs competitively, likely due to effective feature selection and the simplicity of the input space.

**Table 6.**
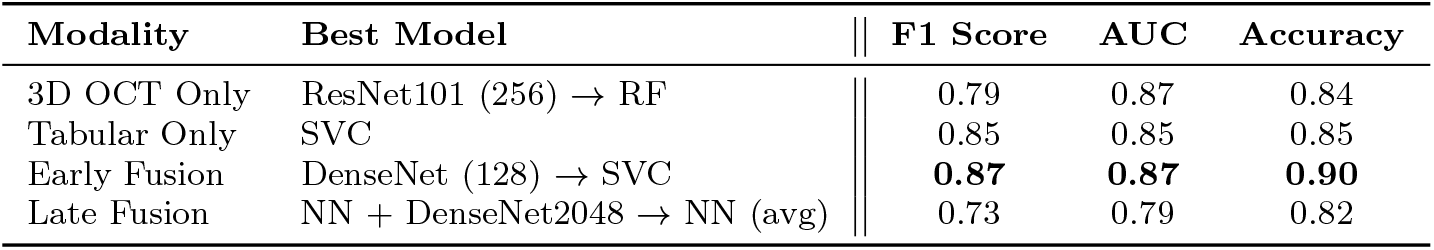
Comparison of best-performing models across unimodal, early fusion, and late fusion strategies. Early fusion with DenseNet and SVC achieves the highest overall performance, while late fusion fails to detect the minority class.

| Modality | Best Model | F1 Score | AUC | Accuracy |
| --- | --- | --- | --- | --- |
| 3D OCT Only | ResNet101 (256) → RF | 0.79 | 0.87 | 0.84 |
| Tabular Only | SVC | 0.85 | 0.85 | 0.85 |
| Early Fusion | DenseNet (128) → SVC | **0.87** | **0.87** | **0.90** |
| Late Fusion | NN + DenseNet2048 → NN (avg) | 0.73 | 0.79 | 0.82 |

## 4 Discussion

In this paper, we performed multimodal ML for pediatric MS classification using OCT, establishing a foundation for future clinical applications in neuroinflammatory disorders. Unlike previous approaches that rely on healthy control groups and often use single-source data, our study addresses a more challenging and realistic diagnostic task, distinguishing MS from other non-inflammatory neurological conditions.

Our best-performing model achieved 90% accuracy in distinguishing MS from these diagnostically complex cases using OCT alone. This was accomplished through an early fusion model that integrates complementary structural features from a single OCT scan. Although both sources derive from the same imaging modality, they represent distinct anatomical and functional domains, enabling the model to capture richer patterns than unimodal approaches. This fusion strategy offers a non-incremental advance, showing that integrating diverse subfield data from the same scan can significantly improve diagnostic performance, particularly in real-world clinical settings where disease boundaries are blurred.

Our prior work demonstrated that ML models applied to OCT scans could distinguish pediatric MS from healthy controls with classification accuracies in the 80% range. [8] While promising, those models were evaluated against control groups lacking neurological pathology. In contrast, this study takes a more clinically rigorous approach, challenging the model to distinguish MS from conditions that may share overlapping features on imaging and presentation. This elevates both the complexity and the practical relevance of the classification task and highlights the added value of a fusion-based architecture for pediatric neurodiagnostics.

While deep learning methods, such as CNNs, have shown impressive results in many medical imaging applications, they often require large, diverse datasets to generalize well. This remains a significant challenge in pediatric research, where cohort sizes are typically small. Prior studies have shown that CNN performance can degrade in such settings due to overfitting and limited representation learning [43–46]. Although our dataset was relatively modest in size, we were able to achieve strong results by using a well-calibrated pipeline that combined OCT features, dimensionality reduction, and early fusion of regionally distinct OCT subfields. This demonstrates that carefully designed ML pipelines can outperform deeper architectures when data is limited.

In the unimodal setting, models trained on 3D OCT scans exhibit strong performance, particularly when using CNN-based feature extractors like ResNet101 and DenseNet in combination with tree-based classifiers. Among these, the combination of ResNet101 (with 256-dimensional embeddings) and Random Forest achieves the highest performance, underscoring the effectiveness of ensemble methods in handling high-dimensional representations. Notably, intermediate embedding dimensions (e.g., 128 or 256) consistently outperform larger ones (for instance, 2048), likely due to reduced risk of overfitting in data-limited scenarios.

In the tabular-only setting, SVC outperforms all other models across evaluation metrics, achieving a balanced sensitivity and specificity that other classifiers often fail to maintain. This suggests that, when features are appropriately selected and scaled, linear classifiers like SVC can offer robust performance in low-dimensional, structured feature spaces. In contrast, models such as neural networks and KNN struggle with class imbalance, frequently misclassifying minority-class instances.

Multimodal integration via early fusion yielded the best overall results. The best model, using DenseNet (128-dimensional embeddings) and SVC, achieved an F1 score of 0.87 and an AUC of 0.90. This demonstrates the advantage of combining imaging and clinical data at the feature level, enabling models to learn richer and more complementary representations. Unlike in the unimodal setting, SVC performs best across most multimodal configurations. This likely results from the compact and well-regularized feature space produced by LASSO, which benefits margin-based classifiers.

Moreover, multiple CNN backbones achieve similarly strong results under early fusion, suggesting that, when paired with effective dimensionality reduction, the specific choice of architecture is less critical. These findings highlight the importance of using fusion strategy and feature representation over backbone selection in multimodal learning.

In contrast, late fusion fails to achieve competitive performance. Although it achieved high accuracy, its F1 score dropped due to the failure to correctly identify minorclass instances. This result indicates that late fusion is poorly suited for imbalanced data and does not effectively capture cross-modal interactions. Unlike early fusion, which facilitates joint representation learning and regularization, late fusion processes modalities independently and lacks mechanisms to resolve inter-modality conflicts or compensate for the weaker performance of individual modalities.

Overall, several key patterns emerged across the unimodal and multimodal experiments. First, compact intermediate representations led to more generalizable and discriminative models, particularly in settings with limited data. This trend is consistent across modalities and fusion strategies. Second, classifier performance was closely linked to the structure of the feature space. SVC excels with low-dimensional, structured inputs, whereas ensemble methods like Random Forest are more effective with high-dimensional embeddings derived from raw volumetric data. Third, early fusion consistently outperformed both unimodal and late fusion approaches, reinforcing the value of joint modeling across modalities. Lastly, while late fusion remains a conceptually attractive strategy, it is highly sensitive to class imbalance and typically underperforms compared to other methods.

Notably, this improved performance is achieved in a realistic clinical setting where the control group includes patients with diverse non-inflammatory conditions, not simply healthy children. This increases diagnostic difficulty and makes the success of our early fusion models even more clinically meaningful.

Despite these encouraging results, this study has limitations. First, the relatively small sample size poses challenges for training deep learning models, which tend to overfit in low-data settings. Although we applied dimensionality reduction, feature selection, and cross-validation to mitigate this issue, certain model configurations may still learn spurious patterns that fail to generalize. Second, class imbalance remains a challenge, as reflected in the disparity between true positive and true negative rates across different models. We used weighted loss functions and balanced sampling strategies to address this, yet sensitivity to the minority class remains limited in some unimodal and late fusion settings.

Future work focuses on several directions to address these limitations and advance model performance. We aim to increase the dataset size to reduce sample scarcity and enhance generalization, especially for underrepresented classes. We also plan to explore advanced fusion techniques, such as attention-based mechanisms and adaptive modality weighting, to better integrate complementary information across modalities. In addition, we investigate deep learning architectures that support end-to-end multimodal learning, which may enable richer cross-modal interactions compared to our current two-stage pipeline. Finally, we will consider incorporating additional clinical biomarkers and demographics to expand the feature space and improve early detection and disease monitoring capabilities.

## Data Availability

The data supporting this study were collected as part of the Neuroinflammatory Registry at the Hospital for Sick Children and are not publicly available due to patient privacy and ethical restrictions. De-identified data may be made available from the corresponding author upon reasonable request and approval by the institutional ethics board. Code used for preprocessing and model development is available from the authors upon request.

## 6 Cometing Interests

S.S., C.C., S.R., and F.K. have nothing to disclose. E.A.Y. has received research support in the last 3 years from the National Multiple Sclerosis Society, Canadian Institutes of Health Research, National Institutes of Health, Ontario Institute of Regenerative Medicine, Stem Cell Network, SickKids Foundation, Peterson Foundation, Multiple Sclerosis Society of Canada, Guthy Jackson Foundation, OMS Life, Canada’s Drug Agency, Garry Hurvitz Chair in Neurology and the Multiple Sclerosis Scientific Research Foundation. She has served on scientific advisory boards for Biogen, Alexion, and Hoffman-LaRoche. DSMB: WCG, IQVA. Co-chief Editor: MS and Related Disorders. Speaker/other Honoraria/Support for Travel: SOPNIA Chile, University of Chile, ECTRIMS, ACTRIMS, Johns Hopkins University, New Brunswick Neurological Society, American Academy of Neurology, Consortium of MS Centers, University of Ottawa, Canadian Institutes of Health Research, Michael Smith Health Research Organization, Medlink. Clinical trials: Alexion, Novartis, Hoffman-LaRoche. Governing Council/Steering Committee: Stem Cell Network, Rare Kids CAN, Cantrain.

## A Diagnoses in the Non-inflammatory Cohort

The non-inflammatory cohort comprised individuals with diverse neurological and systemic conditions. The most frequent diagnosis was Langerhans Cell Histiocytosis (LCH). Other diagnoses included spinal cord infarct, stroke, astrocytoma, recurrent facial palsy without brain lesions, headache with nonspecific white matter (WM) abnormalities, hallucinations, tingling with normal brain MRI, oral ulcers without a neurological diagnosis, functional visual loss with normal exams, nonspecific cord abnormalities, and WM abnormalities including glioma. Several individuals also exhibited nonspecific white matter changes on MRI without a definitive neurological diagnosis.

**Table 1.**
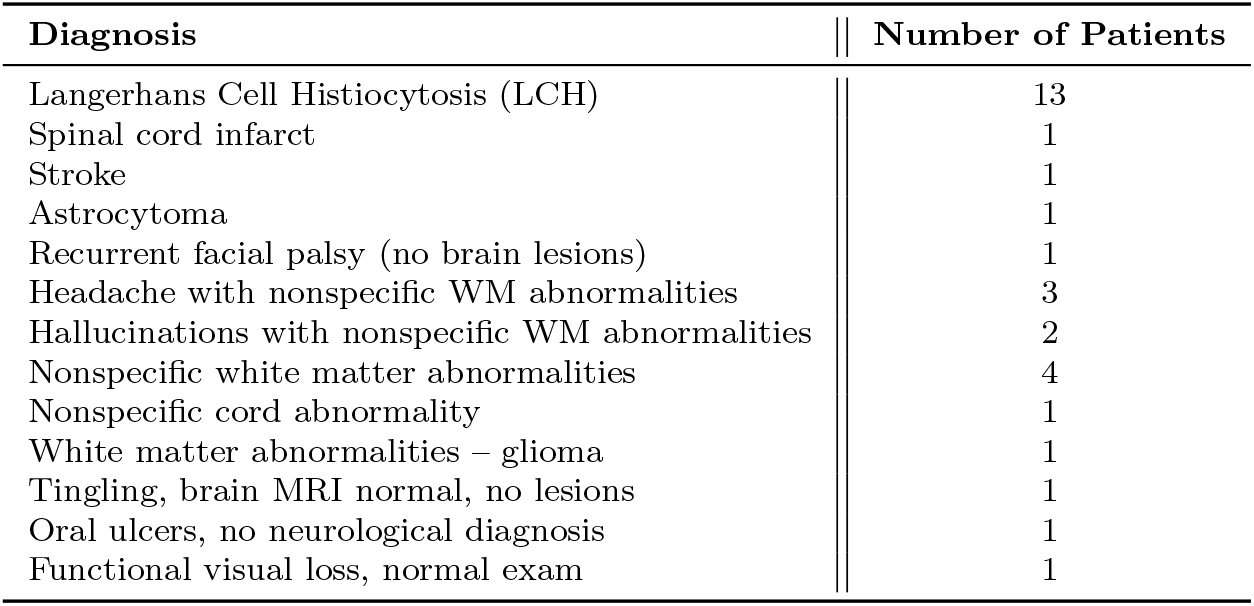
Diagnoses in the non-inflammatory cohort. This table summarizes the alternative neurological and systemic conditions identified among patients without inflammatory demyelinating disease.

## B OCT Features List

**Table 2.**
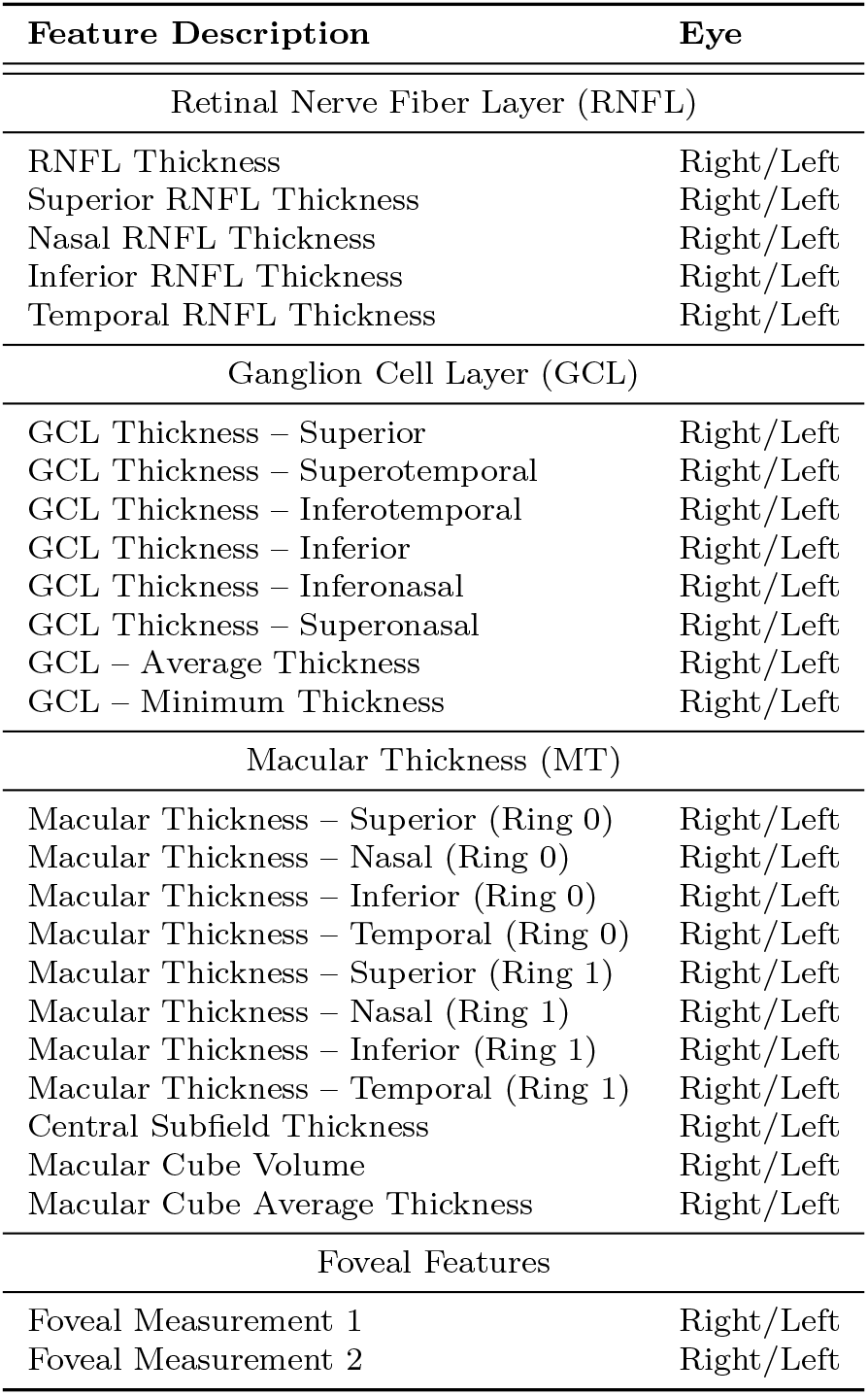
Description of Optical Coherence Tomography (OCT) Features.

## References

[1] Medicine, N.L.: Multiple sclerosis. https://medlineplus.gov/ency/article/000737.htm. Accessed: 2025-05-22 (2025)

[2] Jakimovski, D., Awan, S., Eckert, S.P., Farooq, O., Weinstock-Guttman, B.: Multiple sclerosis in children: differential diagnosis, prognosis, and disease-modifying treatment. CNS drugs 36, 45–59 (2022)

[3] Cerqueira, J.J., Compston, D.A.S., Geraldes, R., Rosa, M.M., Schmierer, K., Thompson, A., Tinelli, M., Palace, J.: Time matters in multiple sclerosis: can early treatment and long-term follow-up ensure everyone benefits from the latest advances in multiple sclerosis. Journal of Neurology, Neurosurgery & Psychiatry (2018)

[4] Yeh, E.A., Graves, J.S., Benson, L.A., Wassmer, E., Waldman, A.: Pediatric optic neuritis. Neurology 87(9 Supplement 2), 53–58 (2016)

[5] Longoni, G., Brown, R.A., Oyefiade, A., Iruthayanathan, R., Wilbur, C., Shams, S., Noguera, A., Grover, S.A., O’Mahony, J., Chung, L., et al.: Progressive retinal changes in pediatric multiple sclerosis. Multiple sclerosis and related disorders 61, 103761 (2022)

[6] Waldman, A.T., Stull, L.B., Galetta, S.L., Balcer, L.J., Liu, G.T.: Pediatric optic neuritis and risk of multiple sclerosis: meta-analysis of observational studies. Journal of American Association for Pediatric Ophthalmology and Strabismus 15(5), 441–446 (2011)

[7] Wexler, M.: Guidelines for MS diagnosis: McDonald criteria. https://multiplesclerosisnewstoday.com/ms-diagnosis-mcdonald-criteria/. Accessed: 2025-05-22 (2023)

[8] Ciftci Kavaklioglu, B., Erdman, L., Goldenberg, A., Kavaklioglu, C., Alexander, C., Oppermann, H.M., Patel, A., Hossain, S., Berenbaum, T., Yau, O., et al.: Machine learning classification of multiple sclerosis in children using optical coherence tomography. Multiple Sclerosis Journal 28(14), 2253–2262 (2022)

[9] Serbecic, N., Beutelspacher, S.C., Geitzenauer, W., Kircher, K., Lassmann, H., Reitner, A., Khan, A., Schmidt-Erfurth, U.: Rnfl thickness in ms-associated acute optic neuritis using sd-oct: critical interpretation and limitations. Acta ophthalmologica 89(5), 451–460 (2011)

[10] Birkeldh, U., Manouchehrinia, A., Hietala, M.A., Hillert, J., Olsson, T., Piehl, F., Kockum, I.S., Brundin, L., Zahavi, O., Wahlberg-Ramsay, M., et al.: The temporal retinal nerve fiber layer thickness is the most important optical coherence tomography estimate in multiple sclerosis. Frontiers in neurology 8, 675 (2017)

[11] Martinez-Lapiscina, E.H., Arnow, S., Wilson, J.A., Saidha, S., Preiningerova, J.L., Oberwahrenbrock, T., Brandt, A.U., Pablo, L.E., Guerrieri, S., Gonzalez, I., et al.: Retinal thickness measured with optical coherence tomography and risk of disability worsening in multiple sclerosis: a cohort study. The Lancet Neurology 15(6), 574–584 (2016)

[12] Yeh, E.A., Marrie, R.A., Reginald, Y.A., Buncic, J.R., Noguera, A.E., O’Mahony, J., Mah, J.K., Banwell, B., Costello, F.: Functional–structural correlations in the afferent visual pathway in pediatric demyelination. Neurology 83(23), 2147–2152 (2014)

[13] Wilbur, C., Reginald, Y.A., Longoni, G., Grover, S.A., Wong, A.M., Mabbott, D.J., Arnold, D.L., Marrie, R.A., Bar-Or, A., Banwell, B., et al.: Early neuroaxonal injury is seen in the acute phase of pediatric optic neuritis. Multiple Sclerosis and Related Disorders 36, 101387 (2019)

[14] Yeh, E., Weinstock-Guttman, B., Lincoff, N., Reynolds, J., Weinstock, A., Madurai, N., Agarwal, N., Buch, P., Karpinski, M., Ramanathan, M.: Retinal nerve fiber thickness in inflammatory demyelinating diseases of childhood onset. Multiple Sclerosis Journal 15(7), 802–810 (2009)

[15] Sajda, P.: Machine learning for detection and diagnosis of disease. Annu. Rev. Biomed. Eng. 8(1), 537–565 (2006)

[16] Shruthi, U., Nagaveni, V., Raghavendra, B.: A review on machine learning classification techniques for plant disease detection. In: 2019 5th International Conference on Advanced Computing & Communication Systems (ICACCS), pp. 281–284 (2019). IEEE

[17] Ahsan, M.M., Luna, S.A., Siddique, Z.: Machine-learning-based disease diagnosis: A comprehensive review. In: Healthcare, vol. 10, p. 541 (2022). MDPI

[18] Varoquaux, G., Cheplygina, V.: Machine learning for medical imaging: methodological failures and recommendations for the future. NPJ digital medicine 5(1), 48 (2022)

[19] Ritter, F., Boskamp, T., Homeyer, A., Laue, H., Schwier, M., Link, F., Peitgen, H.-O.: Medical image analysis. IEEE pulse 2(6), 60–70 (2011)

[20] Shamout, F., Zhu, T., Clifton, D.A.: Machine learning for clinical outcome prediction. IEEE reviews in Biomedical Engineering 14, 116–126 (2020)

[21] Hassan, A.M., Biaggi-Ondina, A., Rajesh, A., Asaad, M., Nelson, J.A., Coert, J.H., Mehrara, B.J., Butler, C.E.: Predicting patient-reported outcomes following surgery using machine learning. The American Surgeon 89(1), 31–35 (2023)

[22] Kumar, D., Pawar, P.P., Gonaygunta, H., Nadella, G.S., Meduri, K., Singh, S.: Machine learning’s role in personalized medicine & treatment optimization. World Journal of Advanced Research and Reviews 21(2), 1675–1686 (2024)

[23] Satheeskumar, R.: Ai-driven diagnostics and personalized treatment planning in oral oncology: Innovations and future directions. Oral Oncology Reports 13, 100704 (2025)

[24] Erickson, B.J., Korfiatis, P., Akkus, Z., Kline, T.L.: Machine learning for medical imaging. radiographics 37(2), 505–515 (2017)

[25] Barragán-Montero, A., Javaid, U., Valdés, G., Nguyen, D., Desbordes, P., Macq, B., Willems, S., Vandewinckele, L., Holmström, M., Löfman, F., et al.: Artificial intelligence and machine learning for medical imaging: A technology review. Physica Medica 83, 242–256 (2021)

[26] Reig, B., Heacock, L., Geras, K.J., Moy, L.: Machine learning in breast mri. Journal of magnetic resonance imaging 52(4), 998–1018 (2020)

[27] Wong, J., Murray Horwitz, M., Zhou, L., Toh, S.: Using machine learning to identify health outcomes from electronic health record data. Current epidemiology reports 5, 331–342 (2018)

[28] Xiao, C., Choi, E., Sun, J.: Opportunities and challenges in developing deep learning models using electronic health records data: a systematic review. Journal of the American Medical Informatics Association 25(10), 1419–1428 (2018)

[29] Gianfrancesco, M.A., Tamang, S., Yazdany, J., Schmajuk, G.: Potential biases in machine learning algorithms using electronic health record data. JAMA internal medicine 178(11), 1544–1547 (2018)

[30] Liu, D.-W., Jia, R.-P., Wang, C.-F., Arunkumar, N., Narasimhan, K., Udayakumar, M., Elamaran, V.: Automated detection of cancerous genomic sequences using genomic signal processing and machine learning. Future Generation Computer Systems 98, 233–237 (2019)

[31] Ren, Y., Chakraborty, T., Doijad, S., Falgenhauer, L., Falgenhauer, J., Goesmann, A., Hauschild, A.-C., Schwengers, O., Heider, D.: Prediction of antimicrobial resistance based on whole-genome sequencing and machine learning. Bioinformatics 38(2), 325–334 (2022)

[32] You, C., Mint, Y., Dai, W., Sekhon, J.S., Staib, L., Duncan, J.S.: Calibrating multi-modal representations: A pursuit of group robustness without annotations. In: Conference on Computer Vision and Pattern Recognition, pp. 26140–26150 (2024).

[33] IEEE Soltanieh, S., Hashemi, J., Etemad, A.: In-distribution and out-of-distribution self-supervised ecg representation learning for arrhythmia detection. IEEE Journal of Biomedical and Health Informatics 28(2), 789–800 (2023)

[34] Chandra, B.S., Sastry, C.S., Jana, S.: Robust heartbeat detection from multimodal data via cnn-based generalizable information fusion. IEEE Transactions on Biomedical Engineering 66(3), 710–717 (2018)

[35] Tewarie, P., Balk, L., Costello, F., Green, A., Martin, R., Schippling, S., Petzold, A.: The oscar-ib consensus criteria for retinal oct quality assessment. PloS one 7(4), 34823 (2012)

[36] Mwanza, J.-C., Oakley, J.D., Budenz, D.L., Chang, R.T., Knight, O.J., Feuer, W.J.: Macular ganglion cell–inner plexiform layer: automated detection and thickness reproducibility with spectral domain–optical coherence tomography in glaucoma. Investigative ophthalmology & visual science 52(11), 8323–8329 (2011)

[37] Breiman, L.: Random forests. Machine learning 45, 5–32 (2001)

[38] Friedman, J.H.: Stochastic gradient boosting. Computational statistics & data analysis 38(4), 367–378 (2002)

[39] Chen, T., Guestrin, C.: Xgboost: A scalable tree boosting system. In: Proceedings of the 22nd Acm Sigkdd International Conference on Knowledge Discovery and Data Mining, pp. 785–794 (2016)

[40] Peterson, L.E.: K-nearest neighbor. Scholarpedia 4(2), 1883 (2009)

[41] Hearst, M.A., Dumais, S.T., Osuna, E., Platt, J., Scholkopf, B.: Support vector machines. IEEE Intelligent Systems and their applications 13(4), 18–28 (1998)

[42] Wang, S.-C.: Artificial neural network. In: Interdisciplinary Computing in Java Programming, pp. 81–100. Springer, ??? (2003)

[43] Raghu, M., Zhang, C., Kleinberg, J., Bengio, S.: Transfusion: Understanding transfer learning for medical imaging. Advances in neural information processing systems 32 (2019)

[44] Soltanieh, S., Etemad, A., Hashemi, J.: Analysis of augmentations for contrastive ecg representation learning. In: 2022 International Joint Conference on Neural Networks (IJCNN), pp. 1–10 (2022). IEEE

[45] Tajbakhsh, N., Shin, J.Y., Gurudu, S.R., Hurst, R.T., Kendall, C.B., Gotway, M.B., Liang, J.: Convolutional neural networks for medical image analysis: Full training or fine tuning? IEEE transactions on medical imaging 35(5), 1299–1312 (2016)

[46] Esteva, A., Kuprel, B., Novoa, R.A., Ko, J., Swetter, S.M., Blau, H.M., Thrun, S.: Dermatologist-level classification of skin cancer with deep neural networks. nature 542(7639), 115–118 (2017)

