## Supplementary Table 1 for "Multimodal Machine Learning for Diagnosis of Multiple Sclerosis Using Optical Coherence Tomography in Pediatric Cases"

### A Diagnoses in the Non-inflammatory Cohort

The non-inflammatory cohort comprised individuals with diverse neurological and systemic conditions. The most frequent diagnosis was Langerhans Cell Histiocytosis (LCH). Other diagnoses included spinal cord infarct, stroke, astrocytoma, recurrent facial palsy without brain lesions, headache with nonspecific white matter (WM) abnormalities, hallucinations, tingling with normal brain MRI, oral ulcers without a neurological diagnosis, functional visual loss with normal exams, nonspecific cord abnormalities, and WM abnormalities including glioma. Several individuals also exhibited nonspecific white matter changes on MRI without a definitive neurological diagnosis.

**Table 1** Diagnoses in the non-inflammatory cohort. This table summarizes the alternative neurological and systemic conditions identified among patients without inflammatory demyelinating disease.

| Diagnosis | Number of Patients |
|---|---|
| Langerhans Cell Histiocytosis (LCH) | 13 |
| Spinal cord infarct | 1 |
| Stroke | 1 |
| Astrocytoma | 1 |
| Recurrent facial palsy (no brain lesions) | 1 |
| Headache with nonspecific WM abnormalities | 3 |
| Hallucinations with nonspecific WM abnormalities | 2 |
| Nonspecific white matter abnormalities | 4 |
| Nonspecific cord abnormality | 1 |
| White matter abnormalities – glioma | 1 |
| Tingling, brain MRI normal, no lesions | 1 |
| Oral ulcers, no neurological diagnosis | 1 |
| Functional visual loss, normal exam | 1 |

### B OCT Features List

**Table 2** Description of Optical Coherence Tomography (OCT) Features

| **Feature Description** | **Eye** |
| --- | --- |
| Retinal Nerve Fiber Layer (RNFL) | |
| RNFL Thickness | Right/Left |
| Superior RNFL Thickness | Right/Left |
| Nasal RNFL Thickness | Right/Left |
| Inferior RNFL Thickness | Right/Left |
| Temporal RNFL Thickness | Right/Left |
| Ganglion Cell Layer (GCL) | |
| GCL Thickness – Superior | Right/Left |
| GCL Thickness – Superotemporal | Right/Left |
| GCL Thickness – Inferotemporal | Right/Left |
| GCL Thickness – Inferior | Right/Left |
| GCL Thickness – Inferonasal | Right/Left |
| GCL Thickness – Superonasal | Right/Left |
| GCL – Average Thickness | Right/Left |
| GCL – Minimum Thickness | Right/Left |
| Macular Thickness (MT) | |
| Macular Thickness – Superior (Ring 0) | Right/Left |
| Macular Thickness – Nasal (Ring 0) | Right/Left |
| Macular Thickness – Inferior (Ring 0) | Right/Left |
| Macular Thickness – Temporal (Ring 0) | Right/Left |
| Macular Thickness – Superior (Ring 1) | Right/Left |
| Macular Thickness – Nasal (Ring 1) | Right/Left |
| Macular Thickness – Inferior (Ring 1) | Right/Left |
| Macular Thickness – Temporal (Ring 1) | Right/Left |
| Central Subfield Thickness | Right/Left |
| Macular Cube Volume | Right/Left |
| Macular Cube Average Thickness | Right/Left |
| Foveal Features | |
| Foveal Measurement 1 | Right/Left |
| Foveal Measurement 2 | Right/Left |
